# Clinical characterization of new-onset chronic musculoskeletal pain in Long COVID: a cross-sectional study

**DOI:** 10.1101/2024.03.09.24304024

**Authors:** Omar Khoja, Bárbara Silva-Passadouro, Elena Cristescu, Katie McEwan, Derek Doherty, Fiona O’Connell, Frederique Ponchel, Matthew Mulvey, Sarah Astill, Ai Lyn Tan, Manoj Sivan

## Abstract

**Purpose:** New-onset chronic musculoskeletal (MSK) pain is one of the common persistent symptoms in Long COVID (LC). This study investigated its clinical characteristics, underlying mechanisms, and impact on function, psychological health, and quality of life.

**Patients and methods:** 30 adults (19 female, 11 male) with LC and new-onset chronic MSK pain underwent clinical examination, Quantitative Sensory Testing (QST), and blood tests for inflammatory markers, and completed the following outcome measures: Timed Up and Go test (TUG), handgrip strength test, COVID-19 Yorkshire Rehabilitation Scale (C19-YRS), Brief Pain Inventory (BPI), Pain Self-Efficacy Questionnaire (PSEQ), Pain Catastrophizing Scale (PCS), International Physical Activity Questionnaire - short form (IPAQ-sf), Generalized Anxiety Disorder (GAD-7), Patient Health Questionnaire (PHQ-9), and EuroQol Five Dimensions health-related quality of life (EQ-5D-5L).

**Results:** New-onset chronic MSK pain was widespread and continuous in nature, and worse in the joints. When compared to normative values reported in the literature: a) QST revealed mechanical hyperalgesia, heightened temporal summation of pain, and hypoesthesia to vibration stimuli, which is strongly suggestive of central sensitization; b) Plasma cytokine assays indicated distinct pro-inflammatory profiles; c) TUG time indicated reduced balance and mobility; d) handgrip strength revealed general weakness; e) physical activity was lower ; and f) there were moderate levels of depression and anxiety with lower self-efficacy scores and lower levels of pain catastrophizing. LC symptoms were of moderate severity (44.8/100), moderate functional disability (22.8/50) and severely compromised overall health (2.6/10) when compared to pre-COVID scores.

**Conclusion:** New-onset chronic MSK pain in LC tends to be widespread, constant, and associated with weakness, reduced function, depression, anxiety, and reduced quality of life. There is associated central sensitization and proinflammatory state in the condition. Further research is essential to explore the longitudinal progression and natural evolution of the new-onset chronic MSK pain in LC.

## 1. Introduction

Long COVID (LC) refers to the persistent symptoms that continue beyond four weeks after an acute COVID-19 infection. LC is a multisystemic condition comprising a wide range of symptoms.^1^ Based on the National Institute for Health and Care Excellence (NICE), LC comprises a) ongoing symptomatic COVID-19 (from 4 to 12 weeks) and b) post-COVID-19 syndrome (beyond 12 weeks).^2^ Conservative estimations anticipate the incidence of LC to be at least 10% of all COVID-19 cases.^3^ This incidence increases up to 30% in moderate non-hospitalized cases and 70% in hospitalized cases.^4^ As of March 2023, the Office of National Statistics estimates that approximately two million people in the UK are experiencing symptoms of LC.^5^ New-onset chronic musculoskeletal (MSK) pain is being increasingly reported as one of the most prevalent and persistent symptoms in LC.^6,7^ Our review comprising 35 observational studies that investigated new-onset MSK pain revealed prevalence rates of up to 65.2%.^8^ This adds to the pre-existing burden of chronic MSK pain before the pandemic, which was estimated to be 1.7 billion worldwide.^9^

Chronic MSK pain is currently among the leading contributor to global disability and affects around a third of the UK population.^10^ The underlying pathophysiological mechanisms associated with chronic pain encompass persistent inflammation that leads to increased sensitivity, disease-related changes in the nervous system, central sensitization, temporal summation, and neuroplastic changes.^11,12^ Psychological and social factors, demographic influences, and genetic predispositions, also contribute to the complexity of chronic pain conditions.

There is a need to understand the presentation and underlying mechanisms of this new chronic pain syndrome in more detail. Existing literature predominantly focuses on documenting the prevalence and location of pain, with only limited attempts to phenotype it and explore its features.^13–17^ A healthy control-matched study revealed that individuals with LC experience heightened pain intensity and interference, severe insomnia, fear of movement, catastrophizing, fear-avoidance beliefs, depression, and anxiety.^15^ In a review aimed to phenotype the post-COVID pain using the 2021 IASP classification criteria, the authors suggested the possibility of nociplastic pain to be the primary mechanism involved in post-COVID pain.^18^ Nociplastic pain is considered difficult to manage and requires a multimodal treatment approach. This was however a hypothesis which needs to be explored in primary research studies.

Understanding the underlying mechanisms of this new pain syndrome will require a diverse range of methods and tools, such as analyzing the cytokine levels and testing the sensory profile of the nervous system. Cytokine storm is considered to be one of the possible mechanisms underlying rapid disease progression in COVID-19.^19,20^ It is also believed that elevation in certain pro-inflammatory cytokines contribute to the initiation and persistence of pathologic pain through direct activation of nociceptive sensory neurons and inflammation-induced central sensitization.^21,22^ Combining the analysis of pro-inflammatory markers with quantitative sensory testing (QST) allows for a multifaceted exploration of pain mechanisms, demonstrating the somatosensory profile of pain and its role in chronicity of pain.^23,24^

This cross-sectional data is derived from the Musculoskeletal Pain in Long COVID (MUSLOC) study, a prospective longitudinal study. It aimed to investigate the clinical characteristics of new-onset chronic pain in LC patients and its impact on physical function, mood, and quality of life. In MUSLOC, we also explored the plausible mechanisms of pain by analyzing the relationship between pain and sensory profile (captured using the QST), and pro-inflammatory blood markers.

## 2. Methods and materials

### 2.1. Study design and Participant selection

We enrolled 30 individuals who met the following inclusion criteria: aged 18 years or older; either tested positive for COVID-19 or reported COVID-19 symptoms that were confirmed by an independent clinician; were clinically diagnosed with LC as defined by NICE guidelines;^25^ were experiencing new-onset MSK pain since COVID-19 infection; had the ability to read and understand English; and were willing to adhere to study procedures. Individuals were excluded if they had pre-existing chronic MSK pain prior to COVID-19 infection.

The primary site of recruitment was the Leeds Long COVID Community Rehabilitation Service. Advertisement flyers on social media, support groups and research websites were secondary methods of recruitment. All individuals who registered an interest to participate were initially contacted by telephone to be assessed for their eligibility to participate in the study. They were appropriately informed about what the study involved, potential risks, disadvantages and benefits of their participation prior to obtaining written informed consent.

This study (clinicaltrials.gov NCT05358119) complies with the ethical principles underlying the Declaration of Helsinki and was approved by London - Central Research Ethics Committee (REC), the Health Research Authority (HRA) and Health and Care Research Wales (HCRW) (Ref 21/PR/1377).

### 2.2. Demographics and Long COVID-related questionnaire

#### Participant demographics

Participants’ demographic variables were captured: age, sex, ethnicity, marital status, height, weight, pre-existing co-morbidities, date of COVID-19 infection, hospitalization due to COVID-19, number and dates of COVID-19 vaccinations, employment category and whether their employment status had been affected by COVID-19 pandemic and LC.

#### COVID-19 Yorkshire Rehabilitation Scale (C19-YRS)

The C19-YRS comprises a Numerical Rating Scale (NRS) of main symptoms of LC (including breathlessness, cough, swallowing, fatigue, continence, pain, cognition, post-traumatic stress disorder (PTSD), anxiety, depression, palpitations, dizziness, weakness, sleep problems, fever and skin rash) and their impact on five daily functions (including communication, mobility, personal-care, activities of daily living, and social role).^7^ Participants were also asked to grade their symptoms prior contracting COVID infection (pre-COVID). The total score of items 1-10 represents the symptoms severity score (0-100), while items 11-15 represent the functional disability score (0-50). The C19-YRS also includes a question capturing the overall health status before and after COVID infection (scores 0-10) in which a score of 0 means the WORST health the respondent can imagine and a score of 10 means the BEST health they can imagine.

### 2.3. Pain evaluation

#### 2.3.1. Clinical assessment

A comprehensive clinical MSK examination assessed the clinical dimensions of MSK pain and explored its characteristics and classification. The clinical examination included an assessment of pain history, duration of pain since onset, frequency (continuous or intermittent), quality of pain (sharp and stabbing, throbbing, aching, dull, burning, numbness and tingling, crushing, and stinging), pain intensity, factors that aggravated and relieved pain, location of pain (upper limbs, lower limbs, chest, spine, and/or widespread), and whether the pain was unilateral or bilateral. Additionally, a pain body map (body chart) was used to mark the exact location of pain. This assessment facilitated the identification of pain type: nociceptive, neuropathic (peripheral/central), nociplastic, or mixed.

#### 2.3.2. Pain-related questionnaires

##### Brief Pain Inventory (Short form) (BPI-sf)

The BPI-sf is a 9-item questionnaire that assesses the location and the severity of pain (worst, least, average, right now), pain medications, amount of relief from pain treatments or medications taken in the last 24 hours, and the interference of pain in an individual’s life (general activity, mood, walking ability, normal work, relationship with other people, sleep, enjoyment of life).^26^ The BPI-sf employs numeric rating scales (NRS) from 0 (no pain) to 10 (pain as bad as you can imagine) for severity, and 0 (does not interfere) to 10 (completely interferes) for pain interference. Total pain severity is the average of the four severity items, and total interference is the average of the seven interference items.

##### The Pain Self-Efficacy Questionnaire (PSEQ)

The Pain Self-Efficacy Questionnaire (PSEQ) is a 10-item questionnaire developed to assess individuals’ beliefs concerning whether or not they are confident to carry out normal activities, despite the presence of pain.^27^ The total score ranges from 0 to 60, a higher score indicating stronger self-efficacy beliefs. Scores above 40 are considered to indicate high levels of self-efficacy, whereas scores below 30 suggest low self-efficacy.^28^

##### The Pain Catastrophizing Scale (PCS)

The PCS assesses the degree of catastrophic cognitions a patient experiences while in pain, in which pain is seen as an extreme threat and the patient suffers exaggerated negative thoughts and feelings around pain.^29,30^ The PCS consists of 13 statements of descriptions of pain experiences and asks individuals to rate the frequency to which they experience these catastrophic cognitions. The total score ranges from 0 to 52, a higher score indicates a high level of pain catastrophizing. A total score of above 30 indicates a clinically significant level of pain catastrophizing.^29^

### 2.4. Quantitative Sensory Testing (QST)

QST was conducted to examine the sensory profile of participants and to evaluate the underlying pain mechanisms. It was performed according to the standardized protocol of the German Research Network on Neuropathic Pain (DFNS).^31^ Participants were instructed to self-report their dominant hand by indicating the hand they most use for various daily activities. Accordingly, QST was done on the palmar side of the participants’ dominant hand. The QST comprised 12 thermal and mechanical parameters testing the function and pathways of the small and large afferent fibers. These parameters include cold detection threshold (CDT); warm detection threshold (WDT); thermal sensory limen (TSL) to assess paradoxical heat sensation (PHS); heat pain threshold (HPT); mechanical detection threshold (MDT); mechanical pain threshold (MPT); mechanical pain sensitivity (MPS); dynamic mechanical allodynia (DMA); wind-up ratio (WUR); vibration detection threshold (VDT); and pressure pain threshold (PPT).

Thermal thresholds were determined using a Q-Sense Thermal Sensory Analyzer (Medoc Ltd., 2012). MDT was assessed using a standardized set of von Frey filaments (OptiHair2 Set, MARSTOCK nervtest, Schriesheim, Germany). MPT was assessed using weighted pinprick stimuli (The PinPrick, MRC Systems GmbH, Heidelberg, Germany). A stimulus-response function for MPS was determined using the same weighted pinprick stimuli as for MPT. Additionally, DMA was tested using light stroking with a cotton wool (Q-Tip) fixed to a plastic strip, a cotton wisp, and a paintbrush. WUR was assessed using repeated skin stimulation with a predefined force and tip size (256 mN weighted pinprick). VDT was assessed with a The Rydel-Seiffer tuning fork (128/64 Hz, 8/8 scale) (US Neurologicals LLC, Poulsbo, WA, United States). The tuning fork was applied at suprathreshold vibration intensity over the participants’ radial styloid process. PPT was assessed using a pressure algometer device (Pain Diagnostic & Thermography (PDT), Italy) with a probe area of 1 cm^2^ that exerts pressure up to 20 kg. The level of pressure when the participant felt the onset of the pain was noted in Kg/cm^2^. The mean of three repeated measurements was calculated, then converted to kPa using the following equation: 1 kg/cm2 = 98.0665 kPa. All participants were examined in a quiet temperature-controlled laboratory (22°C) by the same examiner (OK) following the DFNS protocol testing order. Central sensitization (CS) in participants was recognized if there was one of three specific criteria: abnormally increased MPS, a reduced MPT, or presence of DMA.^32^

### 2.5. Cytokine assays

Blood samples were collected by a phlebotomist from all participants in clotting tubes (BD Vacutainer), then centrifuged (2000 rpm) at 4°C, aliquoted and stored at –80°C. Serum samples were analyzed using two single (IL-17A and C-reactive protein CRP) and a 10-plex panel for proinflammatory cytokines interleukin-1β (IL-1β), IL-2, IL-6, IL-8, IL-10, Interferon-γ (IFN-γ), and tumor necrosis factor-α (TNF-α), using immunoassays from Meso Scale Discovery (MSD, Gaithersburg, MD, USA) following manufacturer’s guidelines. Details of the detection ranges are provided in supplementary material. Analyses were done using a QuickPlex SQ 120 instrument (MSD) and DISCOVERY WORKBENCH^®^ 4.0 software. Cytokine levels were compared to 16 post-infection controls with no LC symptoms (data provided in supplementary material). Levels were dichotomized as raised if above the 95% CI of the distribution of values in no-LC controls, as well as analyzed using linear data.

### 2.6. Physical performance and activity level

#### 2.6.1. Hand-Grip Strength test

Maximal hand strength was determined using a hydraulic hand dynamometer (JAMAR, Sammons Preston, Bolingbrook, Illinois). In a sitting position, with a flexed elbow, forearm neutral, and feet flat on the floor, participants were asked to perform three maximum contractions using the dominant hand (identified by the participant beforehand) each lasting 3–5 seconds and at least 15 seconds apart. The maximum handgrip was determined as the mean of the three measurements.

#### 2.6.2. Timed Up and Go Test (TUG)

Timed Up and Go Test (TUG) was used to measure balance and mobility. Measurement was obtained using a standard armchair (46 cm seat height) and a digital stopwatch (accurate to 0.01 seconds). Participants were seated with their back supported against the chair back. They were instructed to stand up, walk three meters (to a cone on the floor), turn around the cone, walk back to the chair and sit down.^33^ The participants were asked to walk at their normal comfortable pace while wearing their regular footwear, using a walking aid if needed. The TUG was only performed once. The stopwatch was started on the word “go” and stopped as the participant sat down. The TUG time was documented in seconds.

#### 2.6.3. BORG Rating of Perceived Exertion (RPE)

The BORG RPE 6-20 scale was used to assess the effort participant perceived before and after the TUG test had been undertaken.^34,35^ Participants were familiarized with the Borg 6-20 RPE scale before performing the TUG test and were given standardized instructions on how to report their overall feelings of exertion.

#### 2.6.4. International Physical Activity Questionnaire short form (IPAQ-sf)

The IPAQ-sf is a valid and reliable tool to measure the level of physical activity.^36,37^ It consists of seven questions to capture the average daily time spent sitting, walking, and engaging in moderate and vigorous physical activity over the last seven days. Data cleaning and processing were done based on the IPAQ scoring protocol (Guidelines for Data Processing and Analysis of the IPAQ-SF, IPAQ research committee, April 2004). The continuous score, measured in minutes per week, informed the categorical score. This categorization classified participants’ physical activity levels into three categories: Inactive, Minimally active, or HEPA active (health-enhancing physical activity), based on IPAQ guidelines.

### 2.7. Psychological questionnaires and quality of life

#### Patient Health Questionnaire-9 (PHQ-9)

The PHQ-9 is a screening instrument with 9 items, developed and validated to measure depression.^38^ For each item the participants were asked to assess how much they were bothered by the symptoms over the last two weeks. The total score ranges from 0 to 27 and indicates the severity of depression. Scores of 5, 10, 15, and 20 represent cut points for mild, moderate, moderately severe, and severe depression, respectively.

#### General Anxiety Disorder Assessment-7 (GAD-7)

The GAD-7 is a seven-items instrument that is used to measure the severity of generalized anxiety disorder (GAD).^39^ The total score ranges from 0 to 21. Scores of 5, 10, and 15 represent cut points for mild, moderate, and severe anxiety, respectively.

#### EuroQol-5D 5-level (EQ-5D-5L) and EQ-VAS

The EQ-5D-5L is a widely used generic questionnaire to evaluate health-related quality of life, features a five-dimension descriptive system (mobility, self-care, usual activities, pain/discomfort, anxiety/depression) with five response levels, and a Visual Analogue Scale (VAS) where patients rate their health from 0 (worst imaginable health) to 100 (best imaginable health). To drive UK utility values (range from 0 worst health to 1 best health), EQ-5D-5L scores were mapped onto the EQ-5D-3L using Hout et al, mapping crosswalk algorithm.^40^

### 2.8. Statistical analyses

Descriptive statistics were used to summarize variables with mean, standard deviation, minimum and maximum as appropriate for continuous variables, and absolute and relative frequencies and percentages for categorical variables. Paired sample t-test was utilized to assess the variations in C19-YRS symptoms severity, functional disability, and global health scores. It was also applied to determine the significance of the increased Borg score following the TUG test in comparison to the scores before the test. All analyses were performed using MS Excel (version 16.63.1) and IBM Statistical Package for the Social Sciences (SPSS) software version 28.

Data on cytokine levels (linear data) were compared to data in no-fatigue controls using MWU test. Frequency of patients with raised levels were established after determining the 95% confidence interval of the data distribution in no-LC controls. Spearman’s correlation between individual cytokine levels and the pain score were performed (rho and p-values). A Spearman’s correlation-based clustering algorithm (Cluster-3, Stanford University 1998-99) was applied after log transformation, to assess collinearity between cytokines and results were displayed as a heat map using TreeView. Pain score in each patient from the different groups defined by the clustering were compared by ANOVA test.

For QST data, a two-tailed paired t-test assessed differences in BPI intensity score between participants with and without CS signs. Moreover, to compare participants’ QST profiles with a normative dataset from 18 healthy volunteers,^31^ the raw data for each participant were Z-transformed for individual parameters using the following equation:

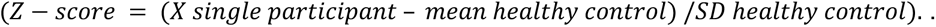

The 95% confidence interval of controls, ranging from −1.96 and +1.96, serves as a benchmark for interpreting the Z-scores. Z-scores above 0 indicate a gain of function, implying that the patients show heightened sensitivity to the tested parameter compared to controls. Whereas Z-scores below 0 indicate a loss of function, signifying decreased sensitivity in patients compared to controls. Values outside the 95% confidence interval and exceeding two standard deviations above or below 0 were considered abnormal. This statistical framework allows for a clear understanding of how patients’ responses differ from the norm, enabling precise assessment of sensitivity changes in the individuals with LC MSK pain.

## 3. Results

### 3.1. Demographic characteristics of participants

Participants had a mean age of 46.8 years (± 11.1) (Range: 26–67 years), 63% were female and the mean Body Mass Index (BMI) was 30.1 (± 11.7). The mean duration from the onset of the MSK pain to the study evaluation was 519.1 days (± 231.7). Only three participants were hospitalized due to COVID-19 infection. Table 1 summarizes the participants’ demographics.

**Table 1:**
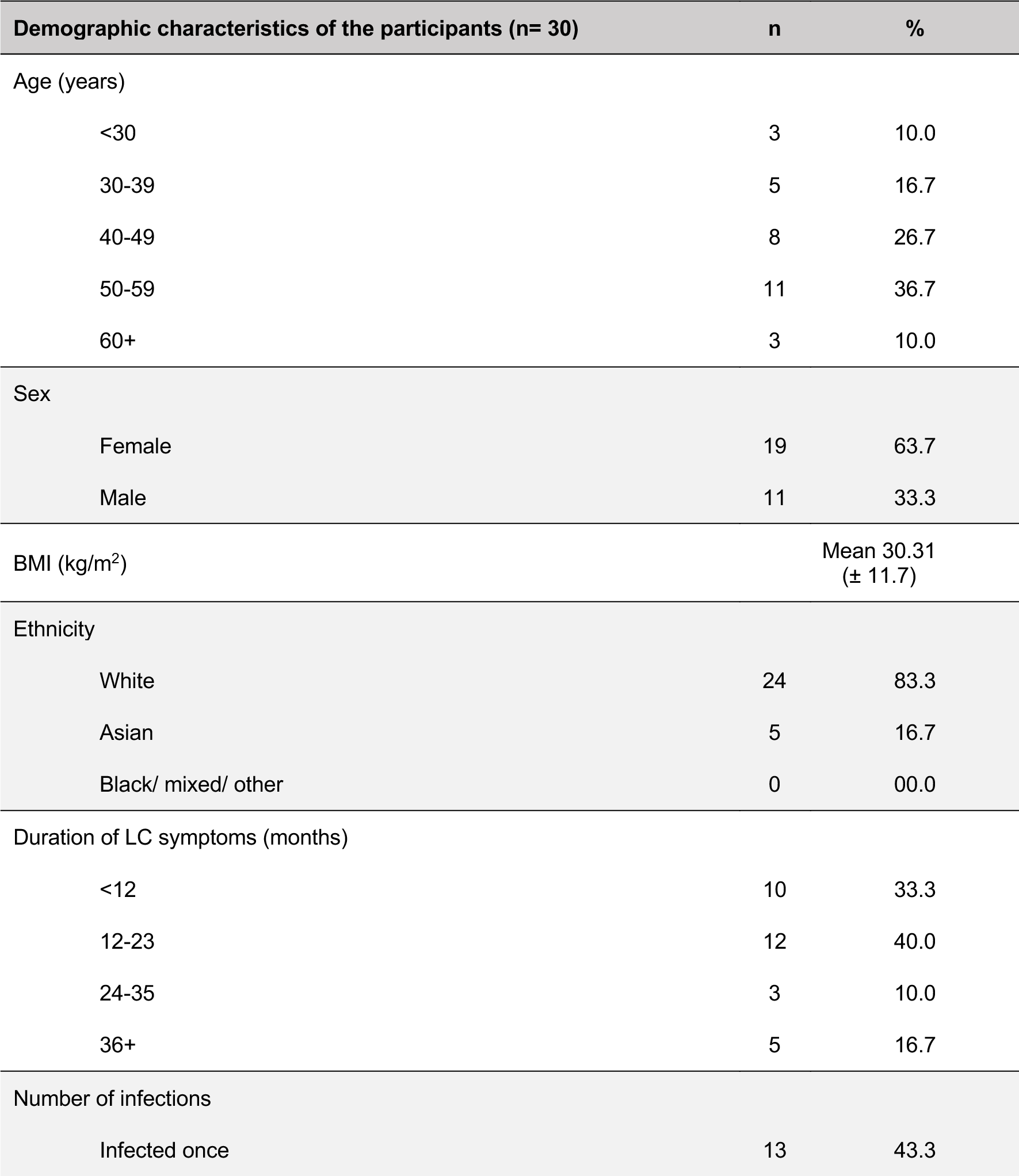
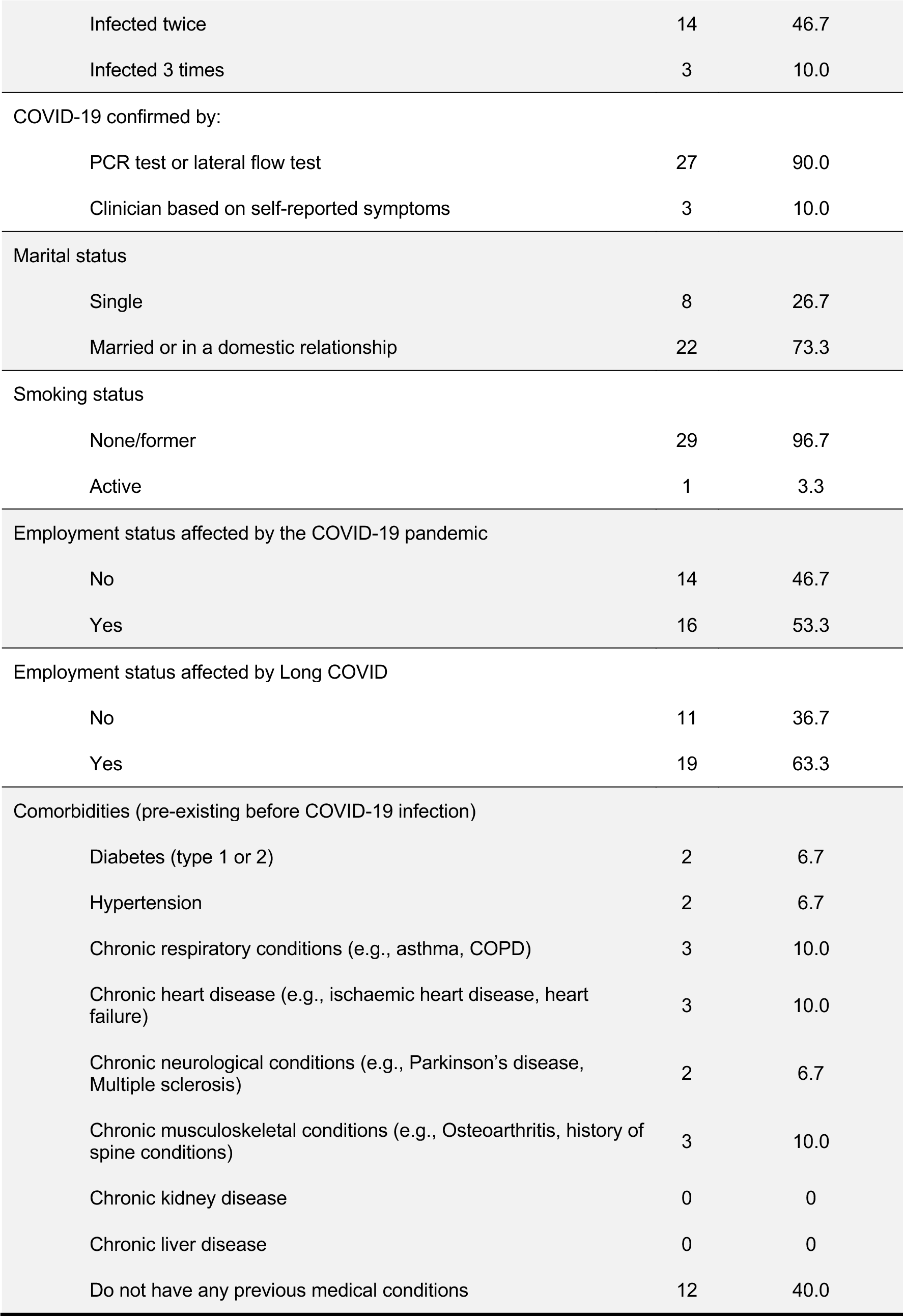
Demographic variables, comorbidities, hospitalization, vaccination, and laboratory confirmation methods.

| Demographic characteristics of the participants (n= 30) |  | n | % |
| --- | --- | --- | --- |
| Age (years) |  |  |  |
|  | <30 | 3 | 10.0 |
|  | 30-39 | 5 | 16.7 |
|  | 40-49 | 8 | 26.7 |
|  | 50-59 | 11 | 36.7 |
|  | 60+ | 3 | 10.0 |
| Sex |  |  |  |
|  | Female | 19 | 63.7 |
|  | Male | 11 | 33.3 |
| BMI (kg/m<sup>2</sup>) |  | Mean 30.31<br>(± 11.7) |  |
| Ethnicity |  |  |  |
|  | White | 24 | 83.3 |
|  | Asian | 5 | 16.7 |
|  | Black/ mixed/ other | 0 | 00.0 |
| Duration of LC symptoms (months) |  |  |  |
|  | <12 | 10 | 33.3 |
|  | 12-23 | 12 | 40.0 |
|  | 24-35 | 3 | 10.0 |
|  | 36+ | 5 | 16.7 |
| Number of infections |  |  |  |
|  | Infected once | 13 | 43.3 |

|  |  |  |
| --- | --- | --- |
| Infected twice | 14 | 46.7 |
| Infected 3 times | 3 | 10.0 |
| COVID-19 confirmed by: |  |  |
| PCR test or lateral flow test | 27 | 90.0 |
| Clinician based on self-reported symptoms | 3 | 10.0 |
| Marital status |  |  |
| Single | 8 | 26.7 |
| Married or in a domestic relationship | 22 | 73.3 |
| Smoking status |  |  |
| None/former | 29 | 96.7 |
| Active | 1 | 3.3 |
| Employment status affected by the COVID-19 pandemic |  |  |
| No | 14 | 46.7 |
| Yes | 16 | 53.3 |
| Employment status affected by Long COVID |  |  |
| No | 11 | 36.7 |
| Yes | 19 | 63.3 |
| Comorbidities (pre-existing before COVID-19 infection) |  |  |
| Diabetes (type 1 or 2) | 2 | 6.7 |
| Hypertension | 2 | 6.7 |
| Chronic respiratory conditions (e.g., asthma, COPD) | 3 | 10.0 |
| Chronic heart disease (e.g., ischaemic heart disease, heart failure) | 3 | 10.0 |
| Chronic neurological conditions (e.g., Parkinson's disease, Multiple sclerosis) | 2 | 6.7 |
| Chronic musculoskeletal conditions (e.g., Osteoarthritis, history of spine conditions) | 3 | 10.0 |
| Chronic kidney disease | 0 | 0 |
| Chronic liver disease | 0 | 0 |
| Do not have any previous medical conditions | 12 | 40.0 |

### 3.2. Long COVID symptoms

The main LC symptoms were captured by the C19-YRS. The participants reported a significant increase in the severity scores for the core primary symptoms of LC compared to pre-infection levels (Table 2). Pain and fatigue were experienced by all participants. Breathlessness was reported by 96% participants, especially during high challenging tasks, such as climbing stairs. Cognition and anxiety were also among the highly reported symptoms, both reported by 92% of the participants.

**Table 2:** Summary of C19-YRS findings.

| C19-YRS items (n=25) | Pre-COVID | Post-COVID |
| --- | --- | --- |
| **Main symptoms of LC** |  |  |
| 1. Breathlessness (% of participants) | 52% | 96% |
| At rest (mean severity, SD) | 0.7 (2.0) | 3.0 (3.3) |
| Dressing (mean severity, SD) | 0.7 (1.5) | 4.3 (3.2) |
| Stairs (mean severity, SD) | 1.0 (1.2) | 6.1 (2.9) |
| 2. Cough/ throat sensitivity/ voice change (% of participants) | 20% | 68% |
| (mean severity, SD) | 0.6 (1.7) | 3.7 (3.4) |
| 3. Swallowing/ nutrition (% of participants) | 8% | 28% |
| (mean severity, SD) | 0.2 (0.8) | 1.5 (2.9) |
| 4. Fatigue (% of participants) | 60% | 100% |
| (mean severity, SD) | 1.3 (1.2) | 8.2 (1.4) |
| 5. Continence (% of participants) | 20% | 48% |
| (mean severity, SD) | 0.4 (1.1) | 2.6 (3.2) |
| 6. Pain/ discomfort (% of participants) | 0% | 100% |
| (mean severity, SD) | 0.0 (0.0) | 6.8 (1.5) |
| 7. Cognition (% of participants) | 48% | 92% |
| (mean severity, SD) | 1.2 (1.6) | 6.0 (2.9) |
| 8. Anxiety (% of participants) | 60% | 92% |
| (mean severity, SD) | 1.7 (1.9) | 5.7 (2.9) |
| 9. Depression (% of participants) | 32% | 68% |
| (mean severity, SD) | 1.2 (2.2) | 4.0 (3.3) |
| 10. PTSD (% of participants) | 20% | 60% |
| (mean severity, SD) | 0.4 (1.0) | 3.2 (3.3) |
| **Symptoms severity score (0-100)** | **7.7 (8.4)** | **44.8 (18.0)** |
| **p Value** | **<0.001** |  |
| 11. Communication (% of participants) | 24% | 72% |
| (mean, SD) | 0.4 (0.8) | 4.2 (3.4) |
| 12. Mobility (% of participants) | 20% | 76% |
| (mean, SD) | 0.4 (1.0) | 4.6 (3.4) |
| 13. Personal-Care (% of participants) | 8% | 52% |
| (mean, SD) | 0.3 (1.2) | 2.1 (2.7) |
| 14. Other Activities of Daily Living (% of participants) | 20% | 92% |
| (mean, SD) | 0.3 (0.7) | 6.6 (2.6) |
| 15. Social role (% of participants) | 12% | 92% |
| (mean, SD) | 0.1 (0.3) | 5.4 (2.8) |
| **Functional disability score (0-50)** | **1.5 (3.0)** | **22.8 (11.3)** |
| **p Value** | **<0.001** |  |
| **Global health (mean, SD)**<br>(0-10) '10 Best Health' | **8.1 (1.2)** | **2.6 (1.4)** |
| **p Value** | **<0.001** |  |

The functional disability score was also increased significantly post-COVID compared to pre-COVID levels. LC symptoms impacted day-to-day living activities in 92% of participants and affected their social role in family care and interactions with friends. Moreover, these symptoms adversely affected most participants’ mobility, communication skills and personal care, resulting in a considerable burden to daily activities compared to the pre-COVID period.

Additionally, participants reported a significant decline in their overall health following the onset of COVID-19, experiencing a drastic decrease from an initial rating of 8.1 (*±*1.2) pre-COVID to a 2.6 (±1.4) out of 10 at the time of assessment. The variation of symptom intensities before and after COVID-19 infection are reported in Table 2. Figure 1 shows symptoms severity score and functional disability score before and after COVID-19 infection.

**Figure 1:**
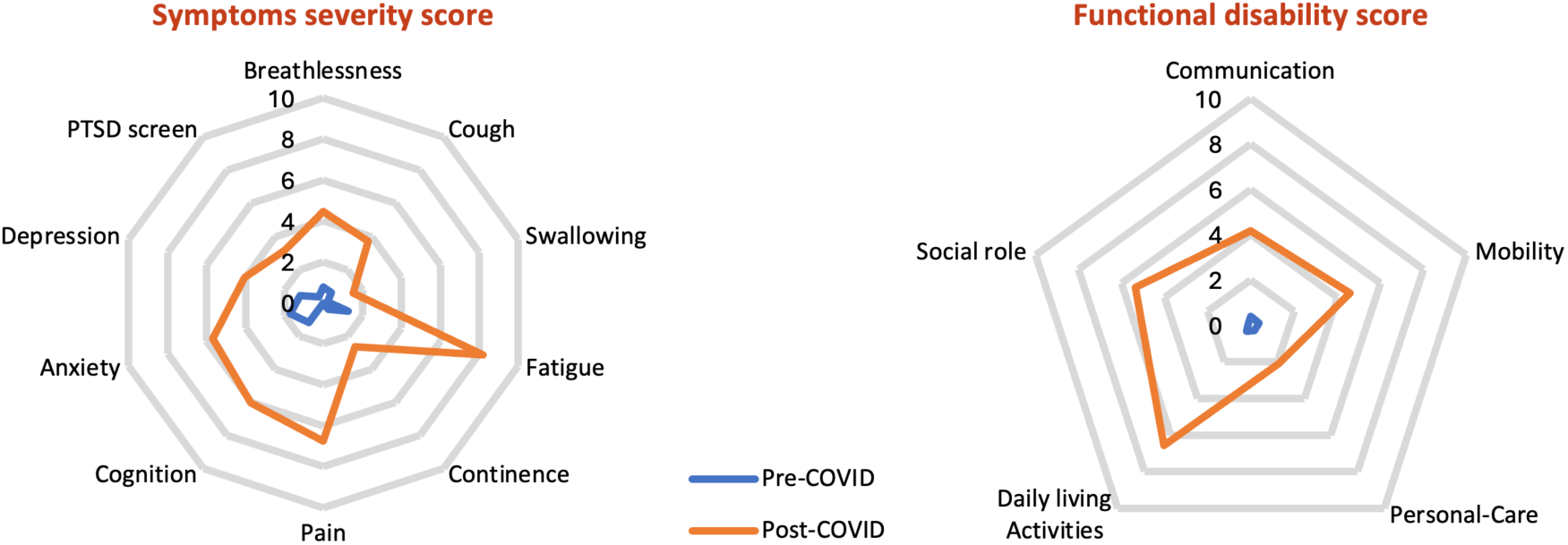
Radar plots of symptoms severity score and functional disability score before (blue radar) and after COVID-19 infection (orange radar). Mild severity < 3, moderate 3-5, severe ≥ 6

### 3.3. Pain characteristics

#### Pain location

The new-onset chronic MSK pain was mostly reported by the participants as generalized widespread pain (90%), characterized predominantly as joint pain: most common sites being the knees (70%), shoulders (63%), cervical spine (60%) and lumbosacral region (57%). Those who did not report having widespread pain (10%), reported experiencing pain in at least four body areas. Figure 2 shows the distribution of pain by location.

**Figure 2:**
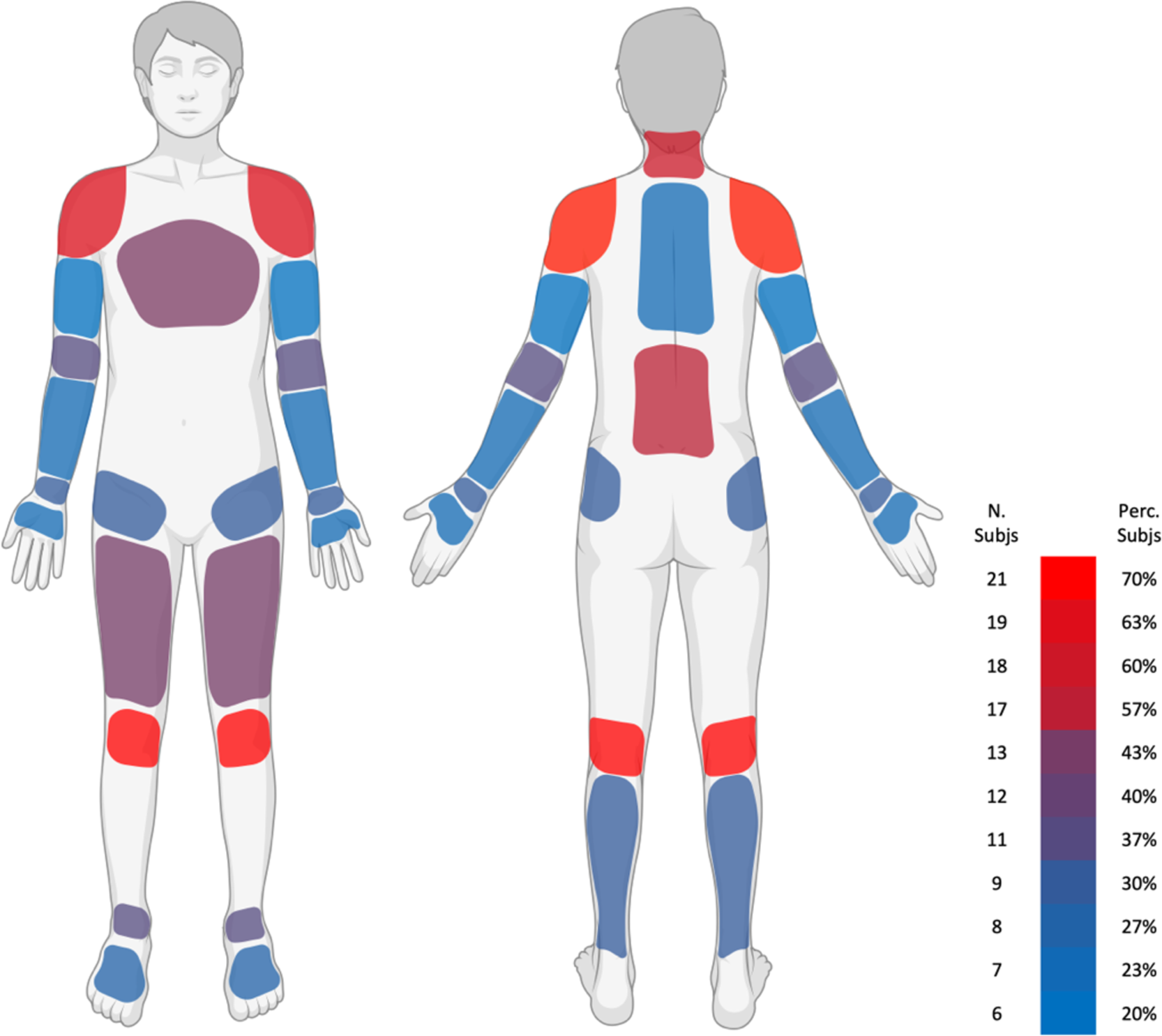
Pain frequency map by location.

#### Pain frequency and quality

90% of the participants were experiencing continuous pain that always remains present, although its intensity may vary throughout the day or from one day to another. The pain quality was mostly described as dull aching pain, with some participants reporting sharp stabbing pain.

#### Pain aggravating and relieving factors

the pain was more likely to be aggravated by excessive activity and fatigue. Resting, sitting, and laying down were the most relieving factors for the pain.

#### Pain intensity and interference

the mean of the BPI total pain severity score was 5.3 (± 1.4), with half of participants (54.2%) falling into the moderate pain category. The mean pain interference in an individual’s life was 6.2 (± 2.2) among our participants, with 84.0% reporting a high interference score. Among the pain interference items, enjoyment of life (mean 7.3 ± 2.4), normal work (mean 6.5 ± 2.5) and mood (mean 6.4 ± 2.8) were the daily functions that were most impacted by pain.

#### Pain self-efficacy

the average score on PSEQ in our study was 25.5 (± 12.6) out of 60. This score indicates the level of self-efficacy our participants feel in managing their pain. A score in this range suggests that the participants might have reduced confidence in their ability to perform daily activities while experiencing pain.

#### Catastrophizing beliefs

the total PCS score among our participants was 20.7 (± 14.2) out of 52. This score, being below the threshold for clinically significant pain catastrophizing, indicates a lower level of catastrophizing within our cohort. In the subcategories, the average scores were as follows: rumination at 7.5, magnification at 3.5, and helplessness at 9.6. These scores provide insight into the specific aspects of catastrophic thinking prevalent in our sample, indicating less negative thought patterns about pain among the participants.

### 3.4. Quantitative Sensory Testing (QST)

#### QST somatosensory profile

In comparison to normative data from Rolke et al,^31^ QST data showed significant gain of function for MPS, which indicates mechanical hyperalgesia, and gain of function for the WUR, suggesting enhanced temporal summation of pain. The data also shows significant loss of function for VDT, indicating hypoesthesia to vibration stimuli, and for the CDT, which suggests hypoesthesia or reduced perception of cold. All other parameters display Z-scores within the 95% confidence interval (Figure 3).

**Figure 3:**
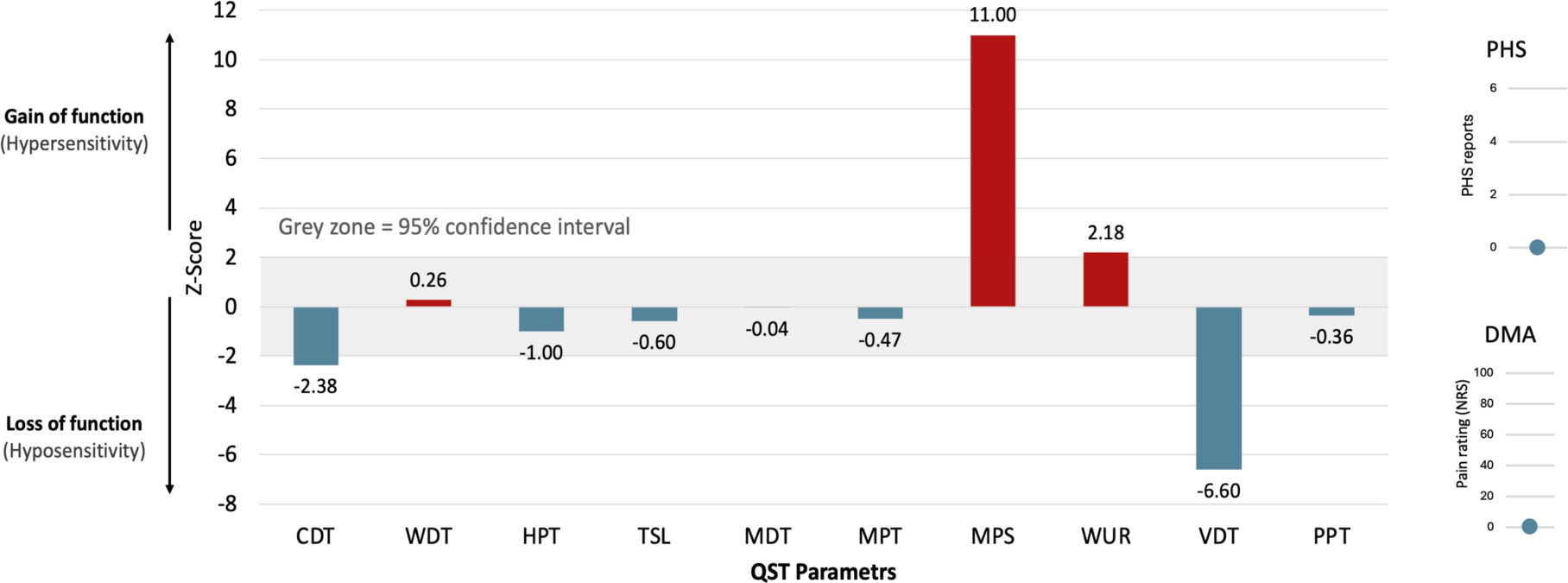
Results of quantitative sensory testing. Z-score = (X single patient − mean healthy controls) / SD healthy controls. CDT, cold detection threshold; WDT, warm detection threshold; HPT, heat pain threshold; TSL, thermal sensory limen; MDT, mechanical detection threshold; MPT, mechanical pain threshold; MPS, mechanical pain sensitivity; WUR, wind-up ratio; VDT, vibration detection threshold; PPT, pressure pain threshold.

#### Central sensitization

In total, 25 participants (83%) showed CS signs. Participants with signs of CS reported significantly higher BPI intensity scores than those without signs of CS, with mean scores of 5.8 (±1.6) and 4.0 (±1.1), respectively (p < 0.05). Overall, QST data revealed mechanical hyperalgesia, heightened temporal summation of pain, and hypoesthesia to vibration stimuli.

### 3.5. Serum cytokine levels

This study’s analyses of inflammatory cytokines revealed a complex relationship with pain. Across our cohort, each participant showed increased levels of at least one cytokine or CRP when compared to controls (Supplementary material), indicating an active inflammation state. The cytokine most frequently increased was IL-13. Of note, only one individual had elevated levels across all tested cytokines. This variability suggested different inflammation profiles among participants. Further investigation into individual cytokine levels using univariate analysis revealed no association between pain scores and any individual cytokines or CRP (Supplementary material).

To define groups of patients with similar cytokine/CRP profiles, we used hierarchical clustering (unsupervised). The analysis, displayed as a heat-map level (Figure 4), showed three distinct distributions of cytokines (group of three cytokines on the left: IFN-γ/TNF-α/IL-10; middle groups of three cytokines: IL-6/IL-8/IL-17 and CRP; and the group on the right: IL-1/IL-2). The analysis also segregated patients into three clusters suggesting that different cytokine profiles can discriminate different patient profiles. Cluster A (five patients) presented a distinct cytokine profile characterized by higher level of IL-10/IFN-γ/TNF-α (displayed in red). Cluster B (twelve patients) displayed only high IL-1β, which potentially indicates a different inflammatory mechanism. Cluster C (thirteen patients) was characterized by high levels of all three groups of cytokines.

**Figure 4:**
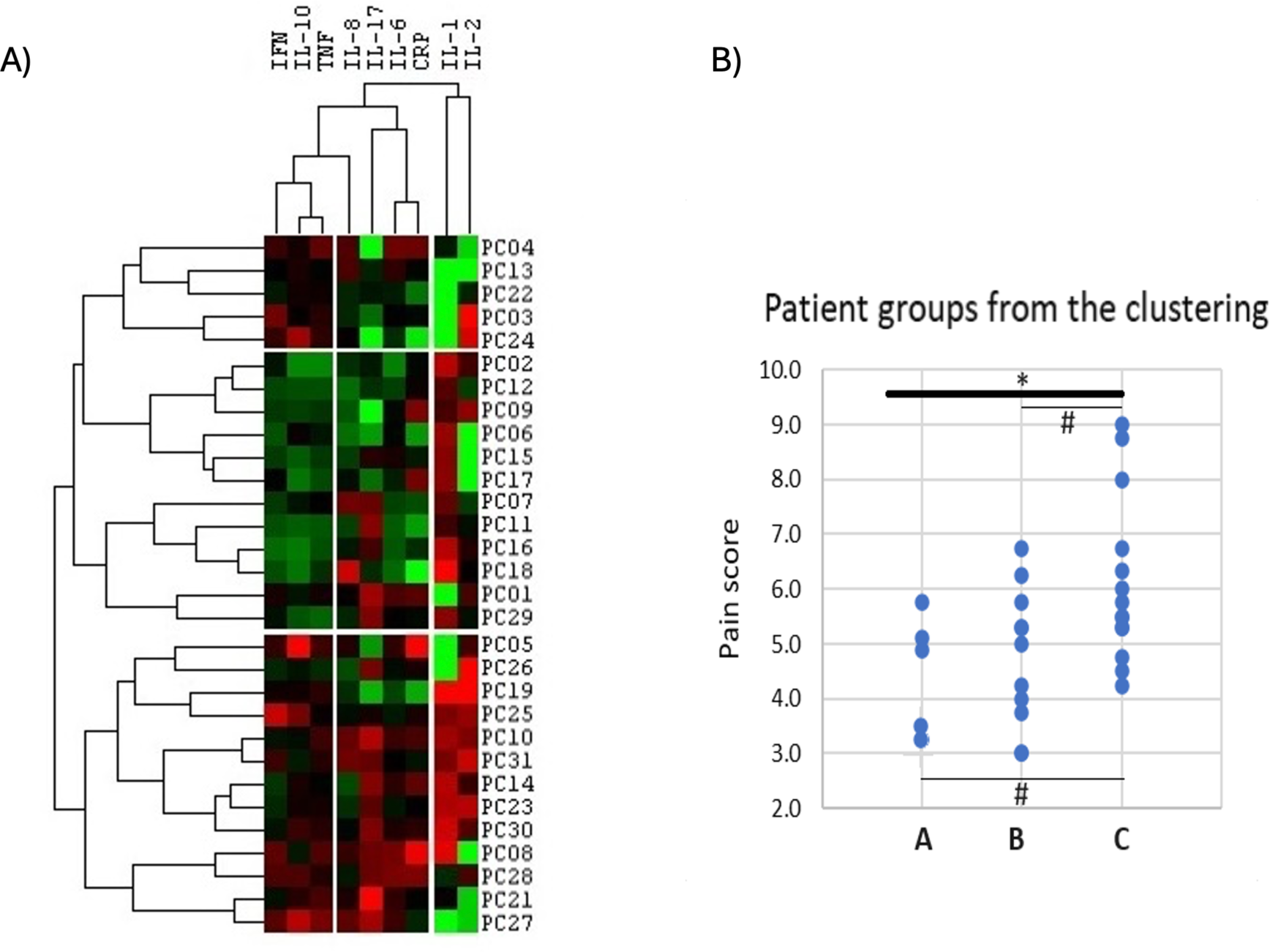
A) Unsupervised hierarchical clustering heat map of cytokines levels. Data are displayed as a heat map with highest levels in bright red and lowest in bright green. Cytokines levels (log transformed) were clustered, defining 3 groups of patients. At the top Cluster-A includes 5 patients with only raised level of IFN-γ/TNF-α/IL-10. In the middle, Cluster-B includes 12 patients with mainly raised levels of most cytokines. B) Scatter plot of pain score across three groups of patients (Cluster A, B and C). There were significant differences in pain scores between the 3 clusters (ANOVA p=0.026) and trend for higher pain score in cluster-C compared to the others.

Pain scores were analyzed in terms of inflammatory patterns as identified in the three clusters of patients (Figure 4, B). Higher pain score was associated with Cluster C, which was characterized by a more pronounced inflammatory profile. Clusters A and B displayed intermediate and lower pain scores, respectively. This data suggests that pain in our study cohort is not the result of isolated cytokine expression but rather a collective pro-inflammatory environment signature.

### 3.6. Functional outcomes

#### Handgrip strength

The overall mean handgrip strength scores of participants’ dominant hands in this study were 21.3 kg (± 6.5) and 10.5 kg (±7.8) for men and women, respectively.

#### TUG test

On average, the TUG time was 13.0 seconds (range 8.93–24.65). 16 participants (53%) showed an elevated risk of falling based on the CDC (STEADI initiative) cut off (> 12 seconds).^41^

#### The Borg RPE

The mean Borg RPE score before the TUG test was 9.2 (± 2.9) and 10.2 (± 2.9) after the TUG test. This reveals a significant increase in perceived exertion scores after the TUG test (t (29) = −4.764, p < 0.001), suggesting that the test induced a higher level of exertion.

#### IPAQ-sf

The self-reported physical activity levels captured by the IPAQ-sf showed overall low activity levels among the participants. According to the IPAQ guideline criteria, 16 (64%) of the participants were categorized as ‘inactive’, and 7 (28%) were classified as ‘Minimally active’. Only 2 (8%) participants reached the criteria of ‘HEPA active’.

### 3.7. Psychological outcomes and quality of life

#### GAD-7

The GAD-7 scores in our study sample ranged from 2 to 20 with a mean score of 10.6 (± 5.1) and with 40% of participants reporting experiencing moderate levels of anxiety. The details of these scores and distribution among participants are presented in Table 4.

**Table 3:** Summary of pain characteristics.

| Outcomes | Values |
| --- | --- |
| **Frequency, n (%)** |  |
| Intermittent (episodic) | 3 (10) |
| Continuous | 27 (90) |
| **Quality, n (%)** |  |
| Dull and aching pain | 28 (93) |
| Sharp and stabbing | 2 (7) |
| **Pain location reported by participants, n (%)** |  |
| Widespread pain | 27 (90) |
| Upper extremity | 27 (90) |
| Lower extremity | 28 (93) |
| Spine | 24 (80) |
| MSK chest pain | 12 (40) |

**Table 4:**
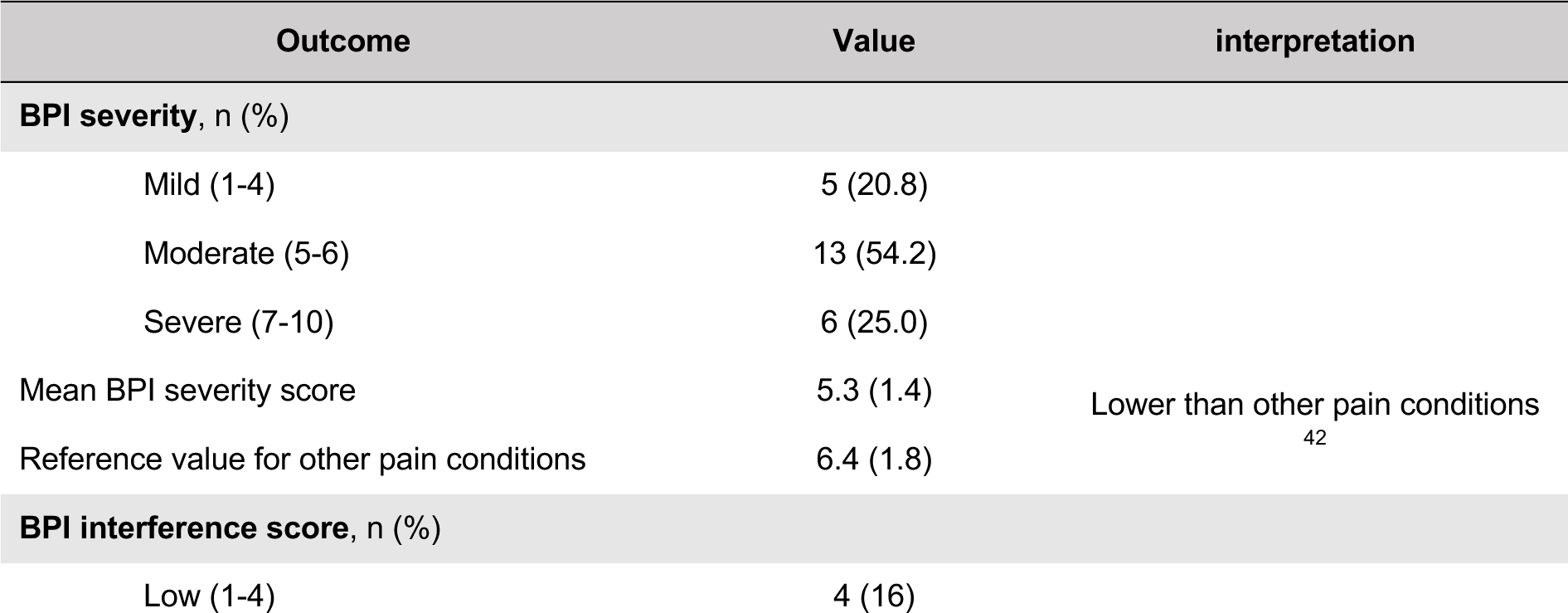
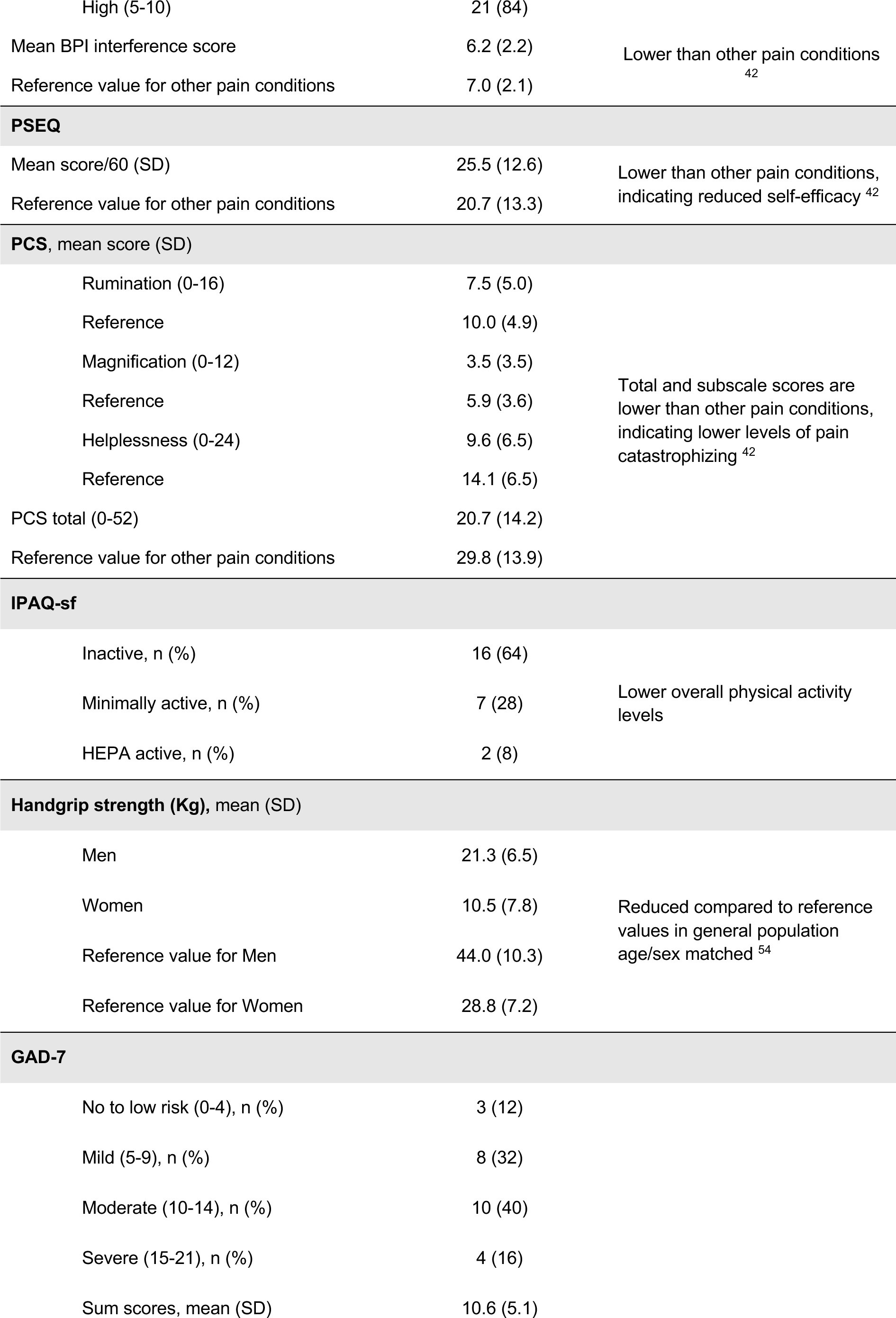
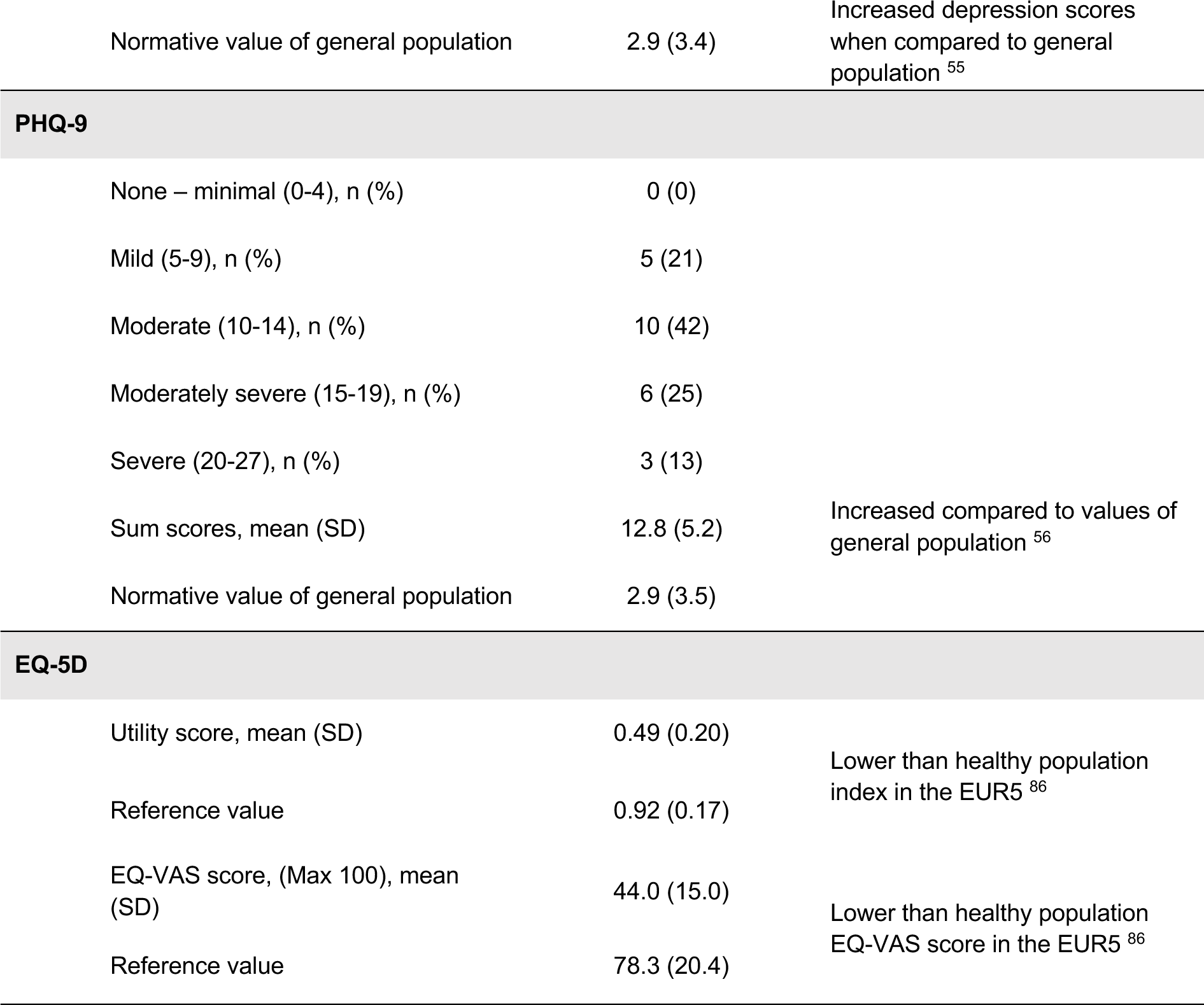
Summary of Key Outcome Measures and Interpretation.

#### PHQ-9

The PHQ-9 scores ranged from 5 to 22, with mean score of 12.8 (± 5.2). This suggests varying levels of depression severity among participants, with 42% of our participants reporting experiencing moderate depression levels, as outlined in Table 4.

#### EQ-5D-5L and EQ-VAS

A significant proportion of the participants reported experiencing moderate problems with activities, pain, and anxiety as measured by the EQ-5D-5L. They generally reported only slight problems with mobility and reported no issues in self-care. The study’s participants had an average EQ-5D-5L index and EQ-VAS scores of 0.49 (± 0.20) and 44.0 (± 15.0), respectively (Table 4). Detailed participant responses across the EQ-5D-5L’s descriptive system are shown in Figure 5.

**Figure 5:**
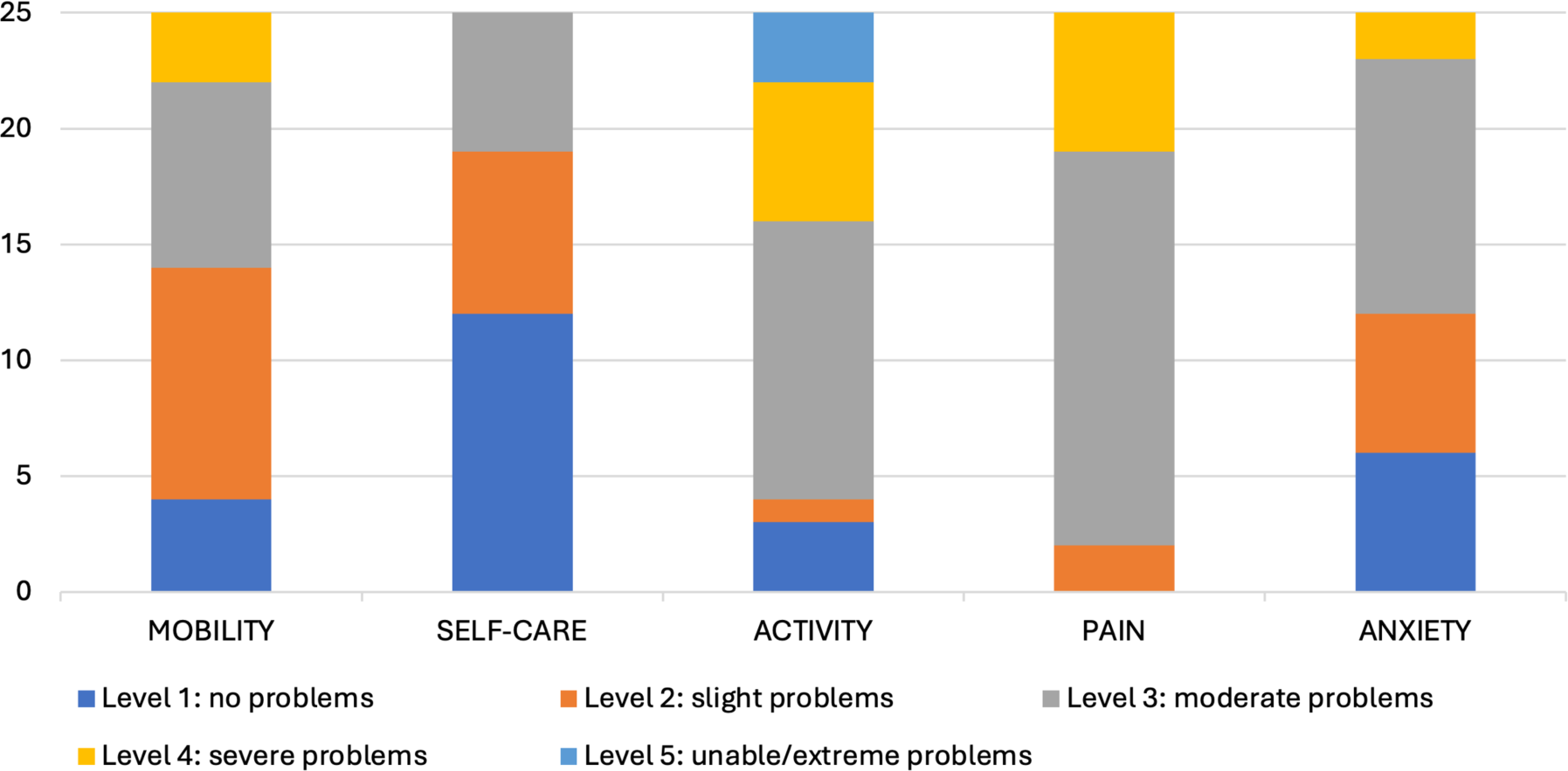
Distribution of responses to the descriptive system of the EQ-5D-5L

## 4. Discussion

In this study, we investigated the clinical characteristics, sensory profile, inflammatory profile, function, mood and quality of life of 30 individuals with new-onset chronic MSK pain in LC. The pain was primarily characterized as generalized, widespread pain that is worse in joints areas of the body. Almost half of the participants experienced a continuous, moderate level of pain that significantly disrupted daily activities. The pain is associated with central sensitization, elevated pro-inflammatory cytokines, weakness, reduced function and physical activity, and depression, anxiety, and reduced quality of life.

The analysis of the BPI revealed that the severity and interference scores of pain in our sample were comparable to those reported in other pain conditions.^42^ In comparison to PSEQ values for individuals with other chronic pain conditions,^42^ the score in our sample falls into a lower percentile. This indicates that our sample had reduced self-efficacy in managing pain relative to other chronic painful conditions.^28^ This lower level of self-efficacy suggests that participants in our study might have decreased confidence in dealing with the ability to perform daily activities despite experiencing pain. Such a finding implies potential challenges in achieving long-term pain management and functional gains.^43,44^ Furthermore, the total score on PCS showed a lower level of catastrophic thinking about pain compared to other chronic painful conditions,^42^ with lower scores in all subcategories of rumination, magnification, and helplessness. This indicates a relatively less negative thought pattern about pain in our cohort (Table 4).

QST data revealed mechanical hyperalgesia and an increased temporal summation of pain, indicative of enhanced small-fiber function, a potential sign of central sensitization.^45^ Additionally, the observed hypoesthesia to vibration stimuli suggests a loss of large-fiber function, aligning with recent findings that COVID-19 is associated with such sensory deficits.^46^ Furthermore, our QST data provide evidence that our participants fulfil the criteria for central sensitization. This is in keeping with evidence in other chronic painful conditions such as Fibromyalgia Syndrome (FMS), Chronic Fatigue Syndrome/Myalgic Encephalomyelitis (CFS/ME), irritable bowel syndrome, and chronic low back pain.^47–49^

Serum cytokine assays indicated a general elevation of pro-inflammatory cytokines and CRP. This persistent inflammatory state may contribute to chronic pain, as inflammation is known to trigger and heighten nerve sensitivity.^50,51^ Levels of individual pro-inflammatory cytokines did not appear to be associated with pain intensity levels, although our sample size is small. Recent reviews suggest that IL-6, IL-10, TNF-α and IFN-y are predictive for LC-related pain in line with our data.^52,53^ Interestingly, in our study, cytokine-clustering grouped IL10, TNF-α, and IFN-y together in the heat-map dendrogram. The different inflammatory clusters of our study suggest that a collective inflammatory signature might be more relevant of pain assessment and management than isolated markers.

Our study showed reduced physical activity, diminished function, low handgrip strength, moderate levels of depression and anxiety, and a decreased health-related quality of life among the participants. Participants exhibited reduced Handgrip strength when compared to age and sex-matched reference values.^54^ Moreover, the GAD-7 and PHQ-9 scores reflected significantly elevated anxiety and depression levels in our cohort, surpassing normative values for the general population.^55,56^ This observation is consistent with recent literature, highlighting pronounced anxiety and depression levels among LC patients.^57,58^ The EQ-5D-5L scores further highlight a considerable disparity in perceived health-related quality of life when compared to the general population in the five largest European economies (EUR5), as detailed in Table 4.

The literature on pain extensively acknowledges that these physical and psychological factors are widely known to worsen the pain experience and increase the probability of pain chronicity.^59,60^ Our results are in strong agreement with the findings from several previous studies for LC.^61–65^ Notably, the observed poor mobility and balance, coupled with lower physical activity and levels of anxiety and depression are recognized as risk factors for increased fall risk.^66,67^ Additionally, these factors are indicative of a heightened risk of mortality and predisposition to other non-communicable diseases.^68^ Such findings underscore the need for integrated interventions targeting physical health and psychological well-being to improve overall health outcomes.

Previous studies on MSK pain in LC have primarily focused on documenting prevalence, with a few studies providing information on its features, characteristics, and location of pain.^15–17,69^ These studies indicated heightened pain intensity and central sensitization in addition to several affected psychological aspects such as fear of movement, anxiety, and depression. Our findings are in agreement with those of Calvache-Mateo et al,^15^ demonstrating heightened pain intensity, central sensitization, depression, and anxiety. This supports the assumptions made by Fernández-de-Las-Peñas et al,^17^ regarding LC MSK pain being a nociplastic type of pain. Our pain location findings are also consistent with Mills et al,^16^ who showed a large proportion of patients reported non-specific MSK pain.

The findings of our study outlined the complexity of the new-onset chronic MSK pain in LC, highlighting the need for a multidimensional approach to treatment. While current guidelines for LC management exist, they lack consensus on treatment specifically addressing chronic pain.^70,71^ A recent umbrella review underscored this gap, noting the absence of pain as a primary outcome in the treatment studies, with the exception of one study that reported pain improvement following Hyperbaric Oxygen Therapy.^71,72^ Emerging intervention studies are evaluating a variety of therapeutic options based on the existing knowledge from similar conditions. These studies are generally investigating the established anti-inflammatory drugs and pain modulators such as nonsteroidal anti-inflammatory drugs and acetaminophen, while also exploring novel therapeutics used in analogous pathologies.^70,73,74^ Nevertheless, the findings of these trials are often limited by validity issues, restricting the applicability to a broader population.^73^ Rehabilitation and non-pharmacological interventions have emerged as safe, applicable, cost-effective, and well-established approach to manage LC symptoms including chronic pain. These interventions focus on improving and restoring the functional ability and quality of life, which align with the multidimensional and complex nature of both LC and chronic pain.^74–76^ Additionally, digital interventions are providing promising avenues to manage chronic pain, physical and mental health impacts of LC, facilitating patient self-management and aid their re-integration into daily life and work, indicating an important step forward in managing LC MSK pain.^76–78^

There are several unique findings in our study which help us understand other related conditions. The widespread nature of new-onset MSK pain has many parallels to FMS and the possible post-viral infection etiology proposed in FMS.^79–81^ Central sensitization and raised cytokines have been reported in FMS and CFS/ME studies.^82^ This also helps us validate some of the conditions which have been poorly understood so far and are associated with considerable social stigma and disbelief, including from healthcare professionals.^83,84^ Our study highlights the need for further research in these chronic pain syndromes to discover biomarkers which can be used as therapeutic targets (for both pharmacological and non-pharmacological treatments).

There are several limitations of this study. The small sample size limits the generalization of findings. We were particularly looking for new-onset pain which made finding suitable patients difficult. Nevertheless, we found 30 participants in one region, which highlights the potential burden of this problem worldwide and how it could add to the existing burden of chronic pain in the populations. The other important limitation of our study is that some outcome measures were based on self-reported data, which introduces recall bias and social desirability bias, potentially affecting the accuracy of participants’ responses. However, subjective reporting is the gold standard extraction method for quantifying pain and psychosocial qualities.^85^ Finally, the abnormal findings in our study although pointing towards plausible mechanisms, cannot be assumed to be causative. These findings are however critical to our understanding of this novel condition which is a public health crisis across the globe.

## 5. Conclusion

Chronic new-onset MSK pain in LC tends to be generalized, widespread, continuous and is associated with central sensitization, elevated pro-inflammatory cytokines, weakness, reduced function and physical activity, depression, anxiety, and reduced quality of life. Future research with larger and more diverse samples will be essential to validate and extend our findings. Furthermore, studies with prospective longitudinal designs would explore the progression and the natural evolution of chronic pain.

## Data Availability

All data produced in the present study are available upon reasonable request to the authors

## 6. Acknowledgments

The authors would like to express their heartfelt gratitude to all participants and their families for their invaluable contribution to this study. Special thanks to the staff of the LCH Long COVID service for their crucial help in recruiting participants. We also acknowledge the support from the Leeds NIHR Biomedical Research Centre and Prof Paul Emery from the Leeds Institute of Rheumatic and Musculoskeletal Medicine.

